# Role of sociodemographic, clinical, behavioral, and molecular factors in precision prevention of type 2 diabetes: a systematic review

**DOI:** 10.1101/2023.05.03.23289433

**Authors:** Dhanasekaran Bodhini, Robert W. Morton, Vanessa Santhakumar, Mariam Nakabuye, Hugo Pomares-Millan, Christoffer Clemmensen, Stephanie L. Fitzpatrick, Marta Guasch-Ferre, James S. Pankow, Mathias Ried-Larsen, Paul W. Franks, ADA/EASD Precision Medicine in Diabetes Initiative, Deirdre K. Tobias, Jordi Merino, Viswanathan Mohan, Ruth J.F. Loos

**Affiliations:** Madras Diabetes Research Foundation, Chennai, India; Department of Pathology & Molecular Medicine, McMaster University, Hamilton, Canada and Population Health Research Institute, Hamilton, Canada; Department of Translation Medicine, Novo Nordisk Foundation, Hellerup, Denmark; Division of Preventive Medicine, Department of Medicine, Brigham and Women’s Hospital and Harvard Medical School, Boston, MA, USA; Novo Nordisk Foundation Center for Basic Metabolic Research, Faculty of Health and Medical Sciences, University of Copenhagen, Denmark; Department of Clinical Sciences, Genetic and Molecular Epidemiology Unit, Lund University, Skåne University Hospital Malmö, Malmö, Sweden; Department of Epidemiology, Geisel School of Medicine at Dartmouth, Hanover, NH, USA; Institute of Health System Science, Feinstein Institutes for Medical Research, Northwell Health, Manhasset, NY, USA; Department of Nutrition, Harvard T.H. Chan School of Public Health, Boston, MA, US; Division of Epidemiology and Community Health, School of Public Health, University of Minnesota; Centre for Physical Activity Research, Rigshospitalet; Institute for Sports and Clinical Biomechanics, University of Southern Denmark; Lund University Diabetes Centre, Department of Clinical Sciences, Lund University, Malmo, Sweden; Oxford Centre for Diabetes, Endocrinology and Metabolism, Radcliffe Department of Medicine, University of Oxford, Oxford, UK; Diabetes Unit and Center for Genomic Medicine, Massachusetts General Hospital, Boston, MA, USA; Dr. Mohan’s Diabetes Specialities Centre, Chennai, India; Charles Bronfman Institute for Personalized Medicine, Icahn School of Medicine at Mount Sinai, New York, NY

## Abstract

The variability in the effectiveness of type 2 diabetes (T2D) preventive interventions highlights the potential to identify the factors that determine treatment responses and those that would benefit the most from a given intervention. We conducted a systematic review to synthesize the evidence to support whether sociodemographic, clinical, behavioral, and molecular characteristics modify the efficacy of dietary or lifestyle interventions to prevent T2D. Among the 80 publications that met our criteria for inclusion, the evidence was low to very low to attribute variability in intervention effectiveness to individual characteristics such as age, sex, BMI, race/ethnicity, socioeconomic status, baseline behavioral factors, or genetic predisposition. We found evidence, albeit low certainty, to support conclusions that those with poorer health status, particularly those with prediabetes at baseline, tend to benefit more from T2D prevention strategies compared to healthier counterparts. Our synthesis highlights the need for purposefully designed clinical trials to inform whether individual factors influence the success of T2D prevention strategies.

## INTRODUCTION

Diabetes affects over 530 million people worldwide.^1^ Around 90% of all diabetes is estimated to be type 2 diabetes (T2D), a non-autoimmune condition with marked pathophysiological heterogeneity.^2^ In many cases, weight loss interventions have demonstrated to delay progression,^3–6^ yet T2D remains a major cause of morbidity and mortality globally.^7^ Chronic inadequate control of hyperglycemia causes downstream microvascular and macrovascular complications that drive the costly and debilitating T2D public health burden.^7^ Coupled with its increasing incidence, public health, and clinical efforts need to optimize effective upstream strategies for T2D prevention.

Landmark randomized intervention trials have demonstrated the effectiveness of intensive lifestyle interventions and glucose-lowering drug therapies for delaying the onset of T2D in patients at high risk.^3–6^ However, T2D incidence has only escalated in the decades since, despite the success of early clinical trials. It is unclear what underscores the underwhelming scalability and translation of effective prevention strategies in real-world settings, and precision prevention research may contribute to understanding this gap.

Precision prevention of T2D serves to minimize an individual’s T2D risk factor profile and maximize the effectiveness of new or established strategies for disease prevention,^8^ through targeting of biological interactions and/or removal of barriers to access and adherence to lifestyle modifications. For example, precision prevention approaches might use clinical (e.g., age, sex, body mass index [BMI]), social (e.g., education attainment, socioeconomic status), or molecular (e.g., genetic, ‘omic traits) characteristics to inform strategies likely to elicit the most effective or sustainable response for an individual, resulting in tailored prevention strategies.^8–10^

The purpose of this systematic review is to critically appraise the accumulated experimental evidence underpinning the feasibility and effectiveness of the clinical translation of precision prevention of T2D. The scope of our investigation included studies reporting the effect modification of lifestyle, dietary, and other behavioral interventions for T2D prevention by any of the following individual-level factors, including sociodemographics, clinical risk factors, behavior, or molecular traits. This work was undertaken as part of a series of systematic reviews conducted by the ADA/EASD Precision Medicine in Diabetes Initiative, an international collaboration of global leaders in precision diabetes medicine.^11^

## METHODS

The systematic review protocol was pre-registered on the International Prospective Register of Systematic Reviews (PROSPERO; CRD42021267686)

### Data sources and search

Our search included MEDLINE, Embase, and Cochrane Central Register of Controlled Trials databases for studies reporting on the efficacy of lifestyle or behavioral interventions with T2D incidence, published from 1/1/2000 to 7/15/2021. An experienced librarian developed a search strategy (**Supplement Table 1**), which included combinations of keywords related to behavioral intervention for preventing T2D (diet, lifestyle, physical activity, body weight), study design, and health outcome, and was limited to the English language. We also scanned the references of included manuscripts and the reference list of systematic reviews published within the past two years to identify additional relevant studies.

### Study Selection

We included studies reporting the effect of a lifestyle, diet, or other behavioral interventions vs. other active comparators or control on the incidence of T2D and reporting the results stratified by any eligible factor. Eligible interventions of interest included lifestyle or behavioral interventions, ranging from education on a single factor to intensive multi-component modification programs, weight loss, diet or supplementation, and physical activity. Eligible stratification factors, or effect modifiers, included individual-level sociodemographic (i.e., race/ethnicity, socioeconomic status/ education, location, age, sex), clinical factors (i.e., BMI, dysglycemia, presence of comorbidities), behavioral (i.e., baseline diet, physical activity) or molecular traits (i.e., genetics, metabolites). We did not review population-level exposures such as built environment, pollution, or climate. Off-label pharmaceutical interventions and bariatric surgery were beyond the scope of the review. We included non-randomized and randomized clinical studies delivering an eligible intervention, comparing against another active intervention, usual care, placebo control or non-control group. We limited inclusion to studies in adults aged >18 years and enrolling at least 100. Studies exclusively among individuals with a current or history of gestational diabetes were excluded because they overlapped in scope with another PMDI consortium review.

### Screening, data extraction, and quality assessment

We used the Covidence online systematic review platform^12^ for literature screening, data extraction, and consensus. Screening consisted of two stages: [1] title and abstract and [2] full text. At each screening stage, two independent reviewers determined the eligibility of the citation, and in the case of disagreement, a third reviewer resolved the discrepancy. Among the full papers accepted for inclusion in the review, two independent reviewers extracted detailed information on the study design, participant characteristics, interventions and comparators, effect modifiers, follow-up for T2D, and analytic approach. We extracted findings related to effect modification of treatment vs. comparator on T2D risk, including strata-specific treatment groups’ T2D cases and incidence rates, or strata-specific treatment-comparator incidence rate ratios, relative risks, risk differences, etc., including measures of variance. We also recorded results for tests for heterogeneity or interaction of the effect modifier with the intervention effect on T2D and noted any text referring to tests performed with “data not shown”. We developed and piloted the data extraction template (**Supplement Table 2**) and discrepancies were ruled on by a third reviewer.

We evaluated the studies’ risk of bias using a modified JBI Critical Appraisal Checklist for randomized controlled trials,^13^ performed by two independent reviewers and disagreements resolved by a third reviewer. We modified the 13-item checklist to 9 questions tailored to evaluating the quality of the study design, but with consideration for our primary interest in stratified results rather than the total intervention effect for T2D risk. These 9 questions were mainly based on randomization, interventions, treatment, and assessor blindness to outcome assessment and our evaluation of these corresponded to color coding in a heat map organized by intervention type and effect modifier (**Supplement Figure 1**).

### Synthesis of results

We collated the literature according to intervention type (e.g., lifestyle programs, dietary supplements) and effect modifier analyzed (e.g., sex, age strata) for the synthesis of results. We determined that a meta-analysis was not feasible among the studies included in our review due to paucity and marked differences in the nature of the study populations, interventions and comparators, and study designs, and effect modifiers analyzed. We qualitatively evaluated the direction and magnitude of results and statistical tests among each prevention strategy for each effect modifier. We weighed these qualitative and quantitative results against their risk of bias and derived the certainty of the collective evidence for an effect modifier’s influence on an intervention strategy, guided by the Diabetes Canada cCinical Practice Scale.^14^ Two reviewers, independently, assessed the certainty of the evidence and resolved disagreements through consensus discussion.

## RESULTS

The results of our systematic literature search are presented in the **Figure 1** attrition diagram. Of the 10,880 citations identified through database searches and other sources, 1,047 abstracts were retrieved for full-text review. From these, 80 publications met our inclusion criteria and data were extracted.

**Figure 1:**
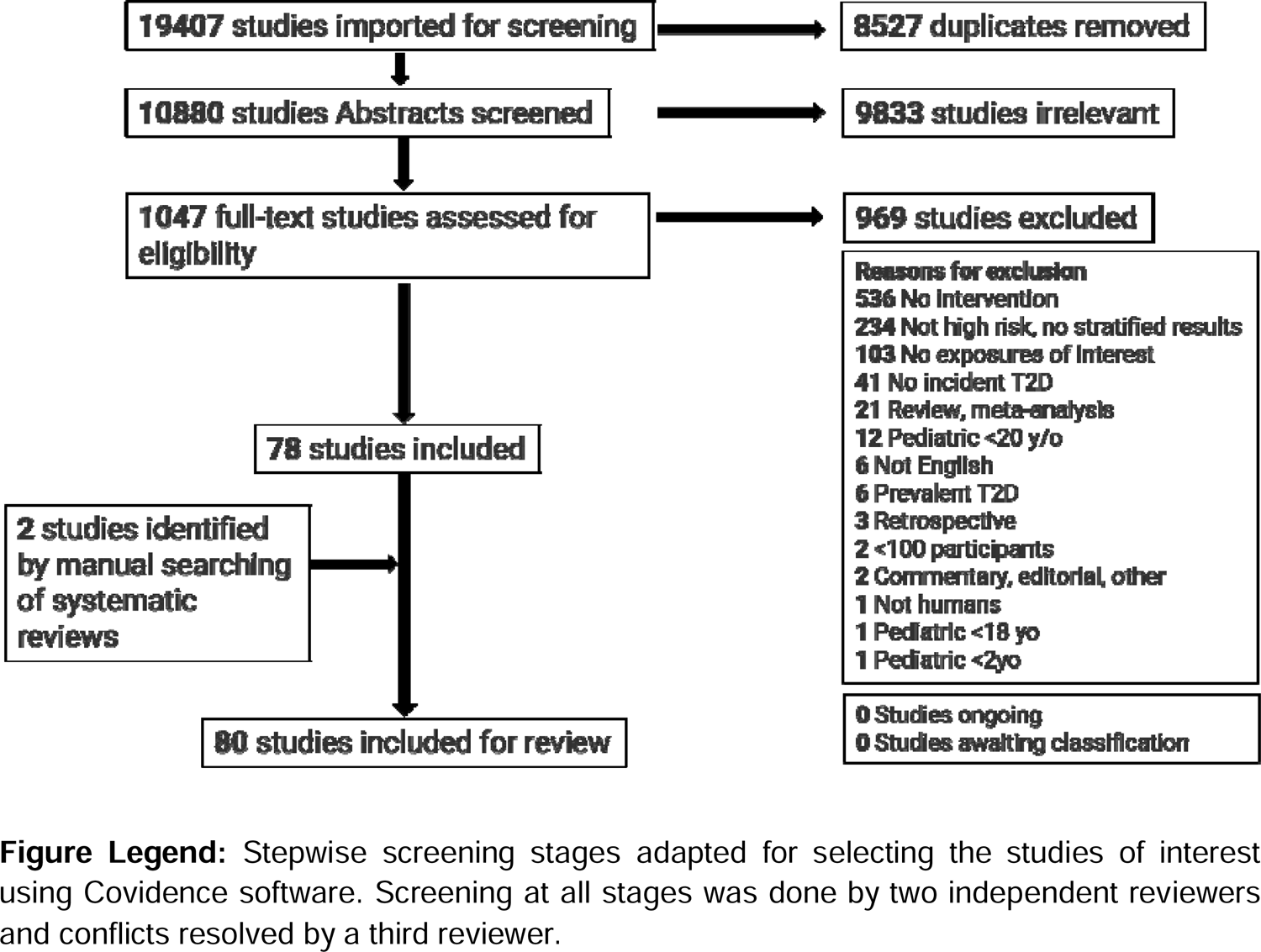
Study Screening Attrition Diagram

### Study characteristics

The 80 publications included in our review represented 33 unique intervention studies (**Table 1, Supplement Table 3**). Twenty-eight studies were randomized clinical trials (RCTs), three studies were nonrandomized parallel group trials, and two studies were single-arm clinical interventions. Fourteen intervention studies took place in Asia, 11 in Europe, seven in North America, and one was a multicenter study that took place in Asia and Europe. Intervention enrollment sample sizes ranged from 302 to 48,835 participants (**Table 1).** Twenty-two studies included individuals at high risk for T2D, two studies at increased cardiovascular risk, and other studies included the general population or other specific groups. The active intervention times ranged from one single lifestyle counseling visit to active interventions lasting up to 10 years (**Supplement Figure 2)**.

**Table 1:**
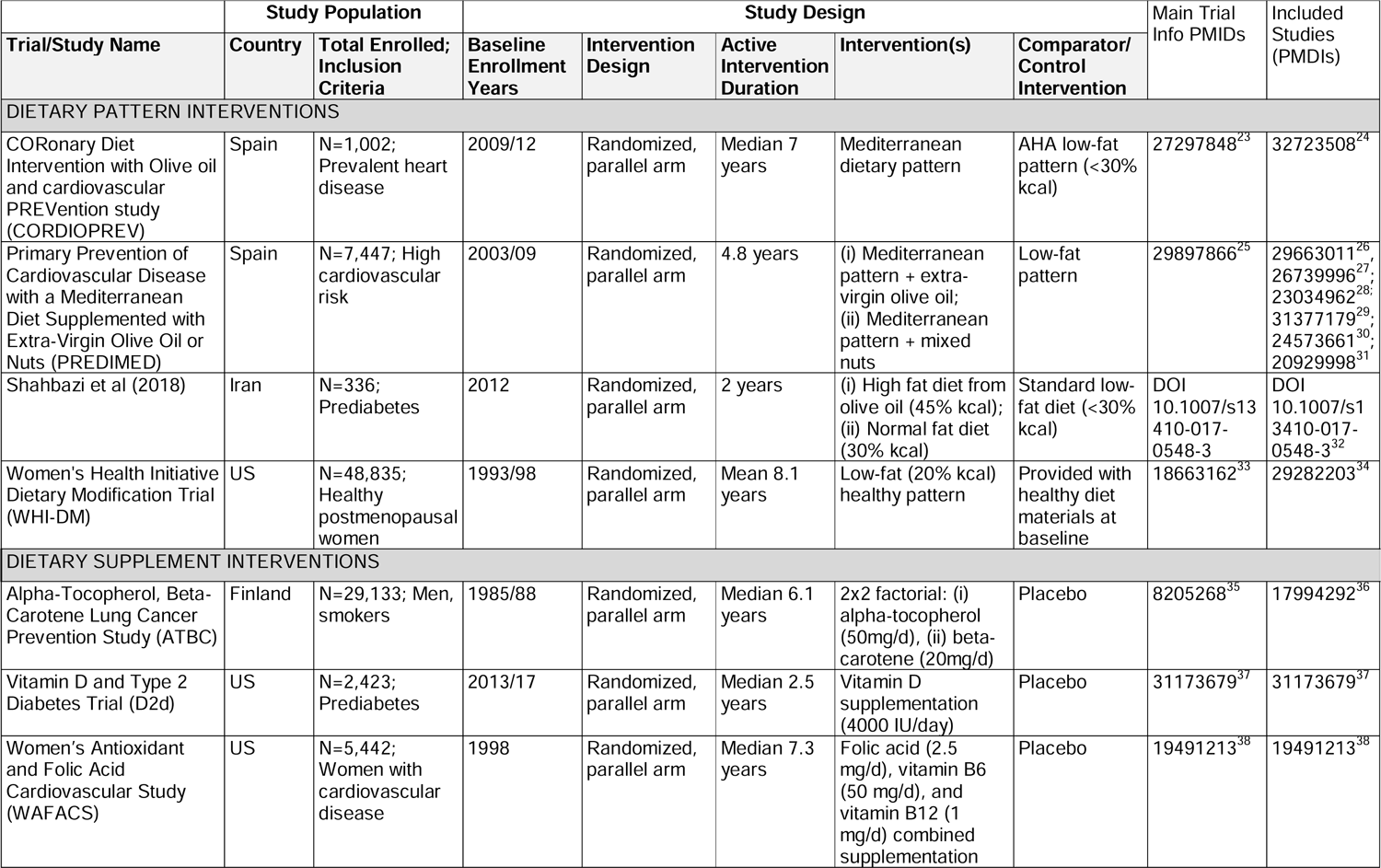
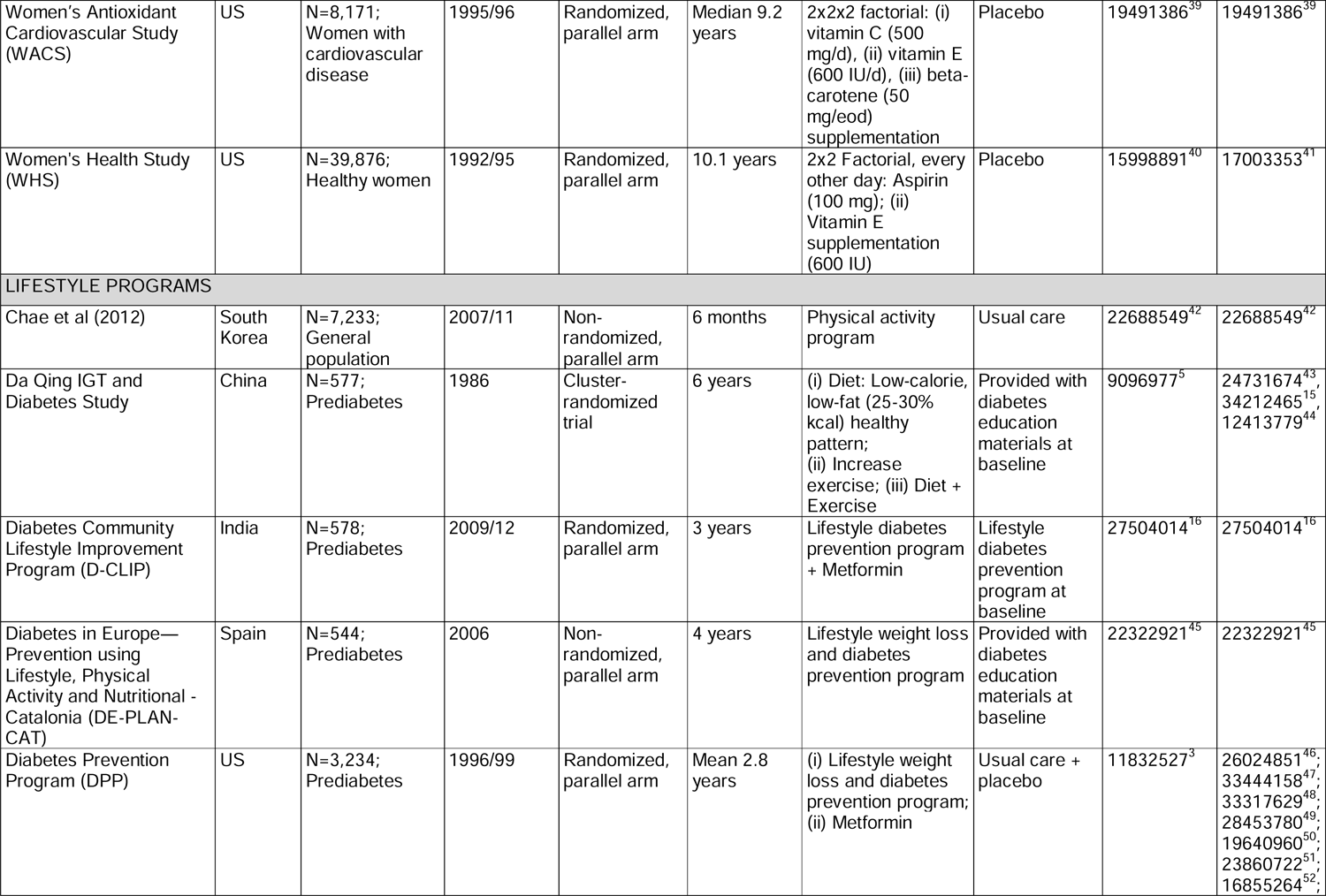
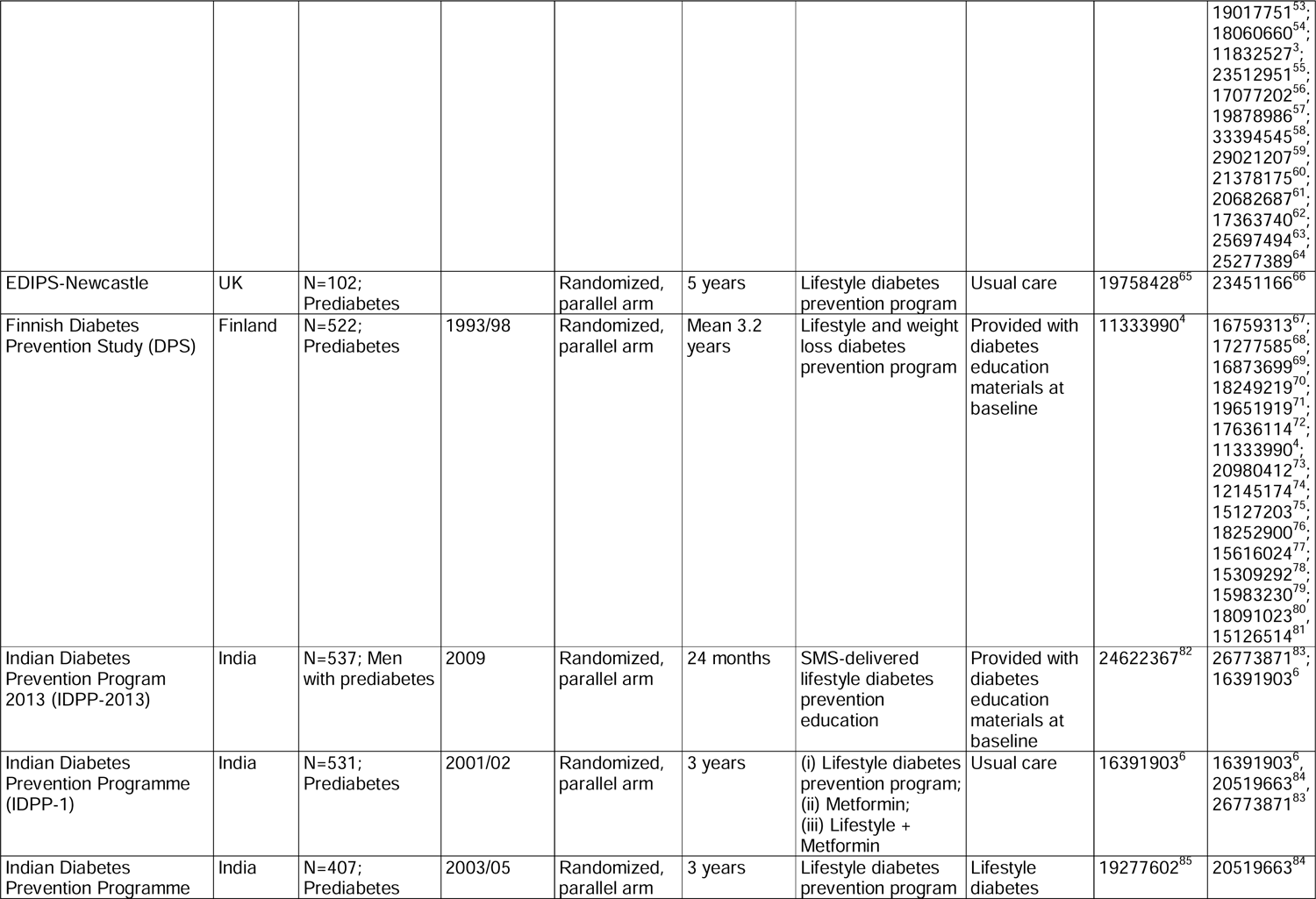
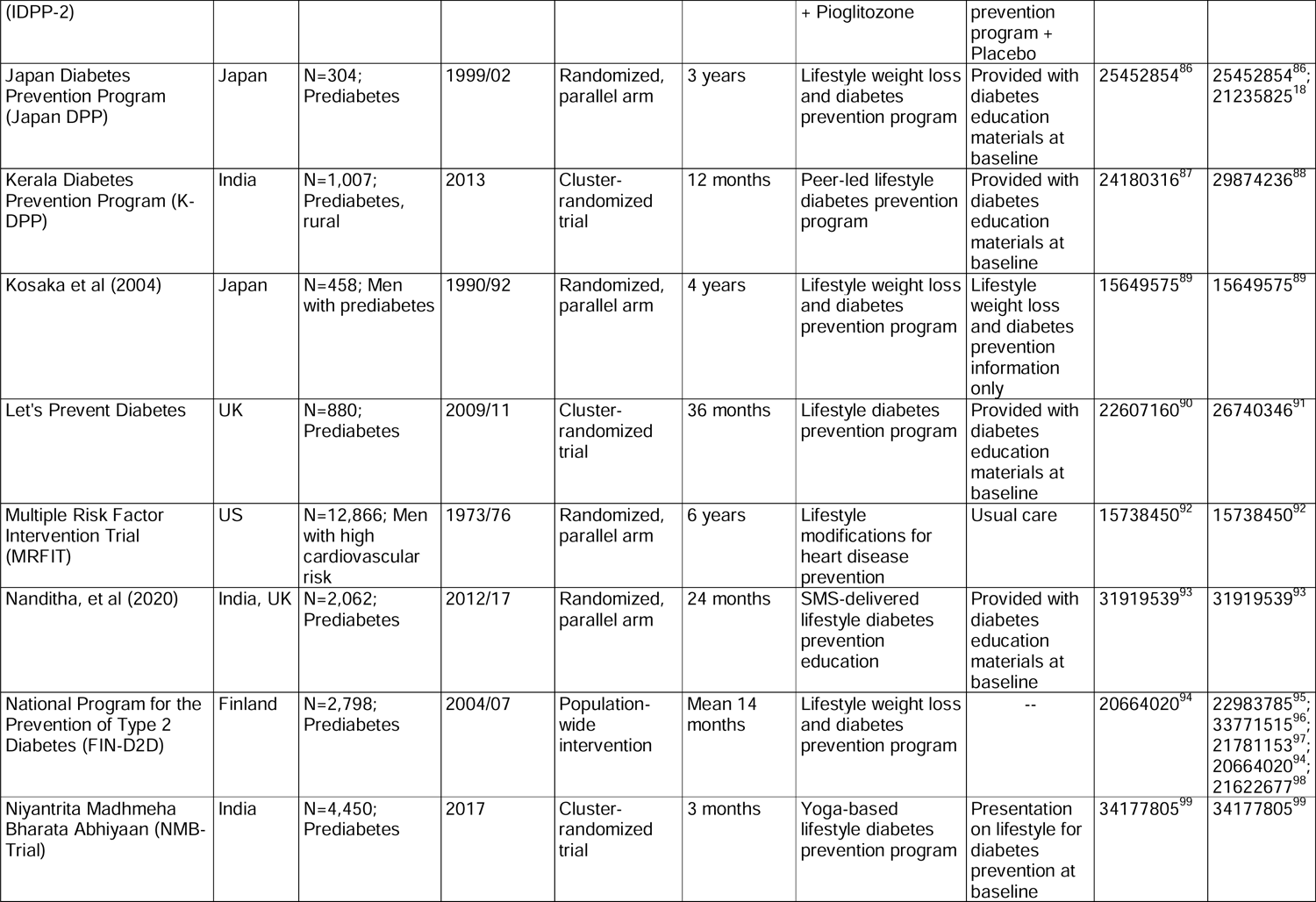
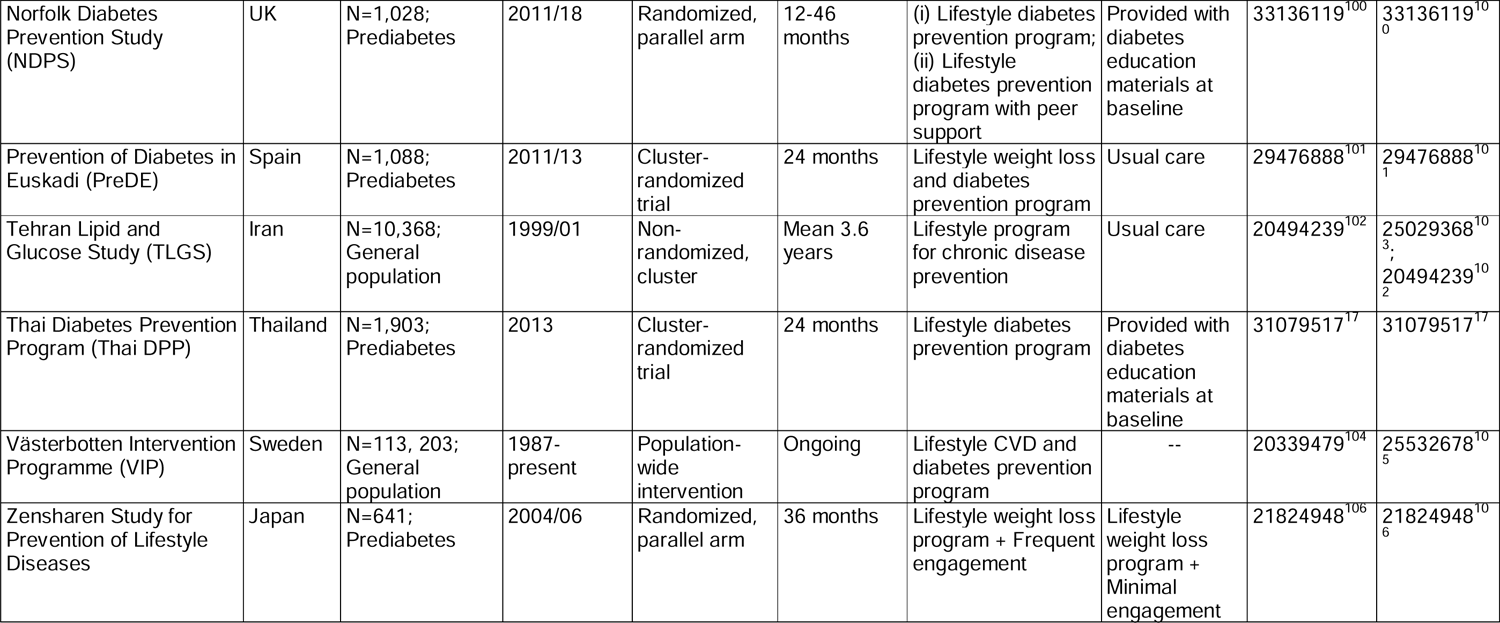
Description of study population and study design of the included trials grouped according to the type of intervention

Twenty-four of the included studies assessed the effect of a multi-component lifestyle intervention program, focused on changes in diet, physical activity, smoking, or body weight loss. Four studies implemented a dietary intervention, and five administered supplements. Across multi-component lifestyle intervention studies, the comparator consisted of a less intensive lifestyle program consisting of usual care or general lifestyle advice administered at baseline. Active comparator groups for dietary intervention studies that focused on high-fat diets consisted of a low-fat intervention. The active comparator for supplement studies consisted of a placebo intervention. T2D was diagnosed in-person with an oral glucose tolerance test (OGTT) in 27 studies, whereas in 6 studies, T2D was ascertained via self-report or through linkage with a healthcare registry database. The primary endpoint was T2D incidence in 21 studies or a composite cardiovascular event in six studies (**Table 1, Supplement Table 3**).

All except seven studies consisting of a multi-component lifestyle intervention program showed evidence that a lifestyle intervention reduces the risk of T2D, with estimated relative risk reduction ranging from 60% to 23% (**Supplement Table 3**). Available evidence also suggests that a high-fat diet, compared to a low-fat diet, reduces the relative risk of T2D. Evidence from studies using supplements showed a null effect on T2D risk reduction.

Our risk of bias assessment determined that the primary study design and approach was generally low, particularly for the RCTs, owing to randomization methods and uniform outcome assessment (**Supplement Figure 1**).However, common concerns for bias were due to non-blinding of participants, deliverers, and outcomes assessors to treatment assignment. Nonrandomized interventions and RCTs having additional concerns for study design did have ratings of high risk of bias.

### Sociodemographic and clinical factors

Some clinical trials, such as the Diabetes Prevention Program (DPP), the Finnish Diabetes Prevention Study (DPS), or the PREDIMED study, were highly represented, with 20, 16, and 6 different publications from each study, respectively. Evidence presented in studies investigating the effect of a lifestyle intervention according to differences in sociodemographic and clinical characteristics did not indicate statistically different effects for age, sex, race/ethnicity, or socioeconomic status. Evidence from studies investigating sociodemographic interaction effects in dietary modification or supplementation trials showed no significant heterogeneity in response to intervention according to these characteristics (**Table 2**, **Figure 2**).

**Figure 2:**
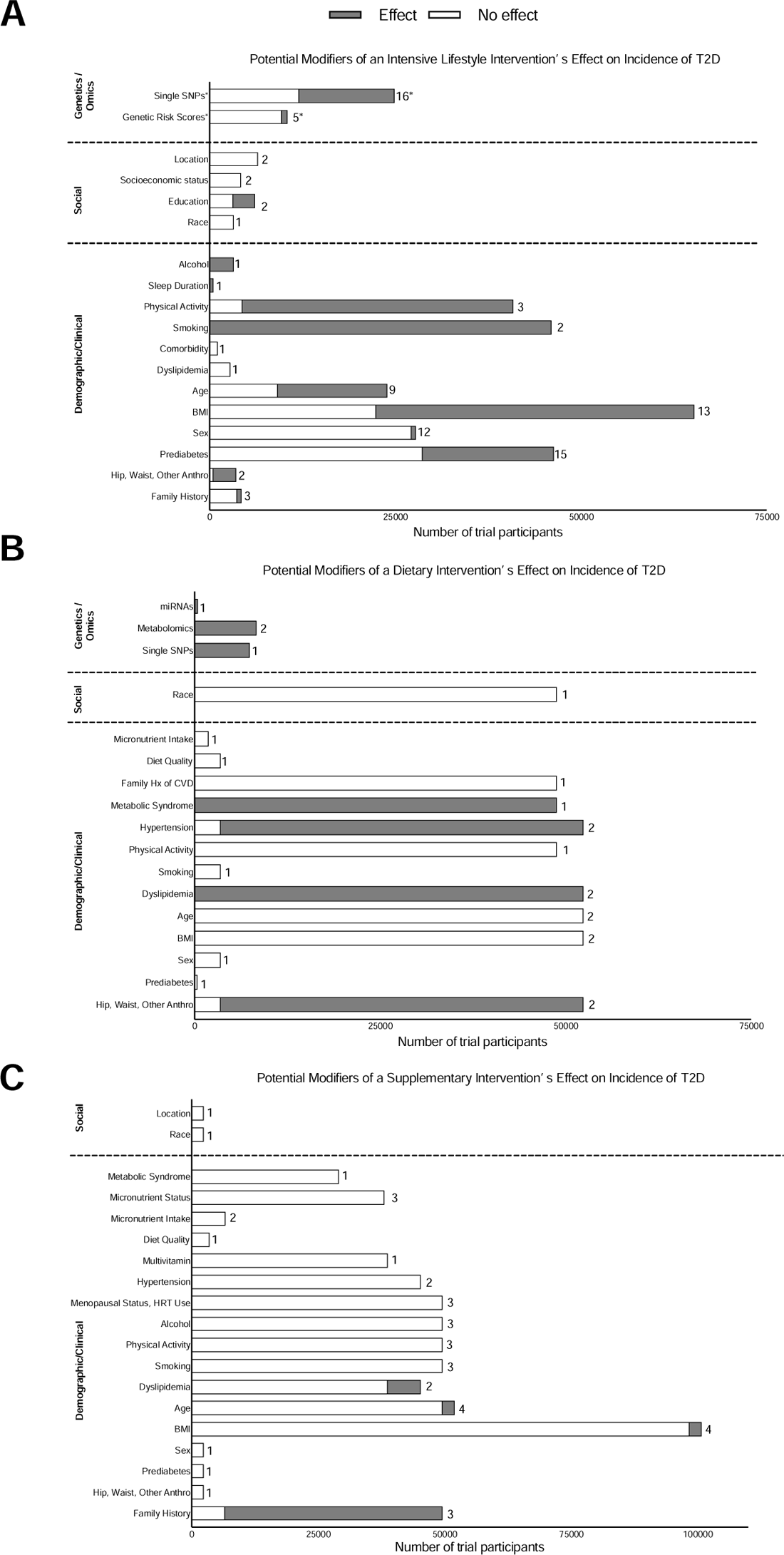
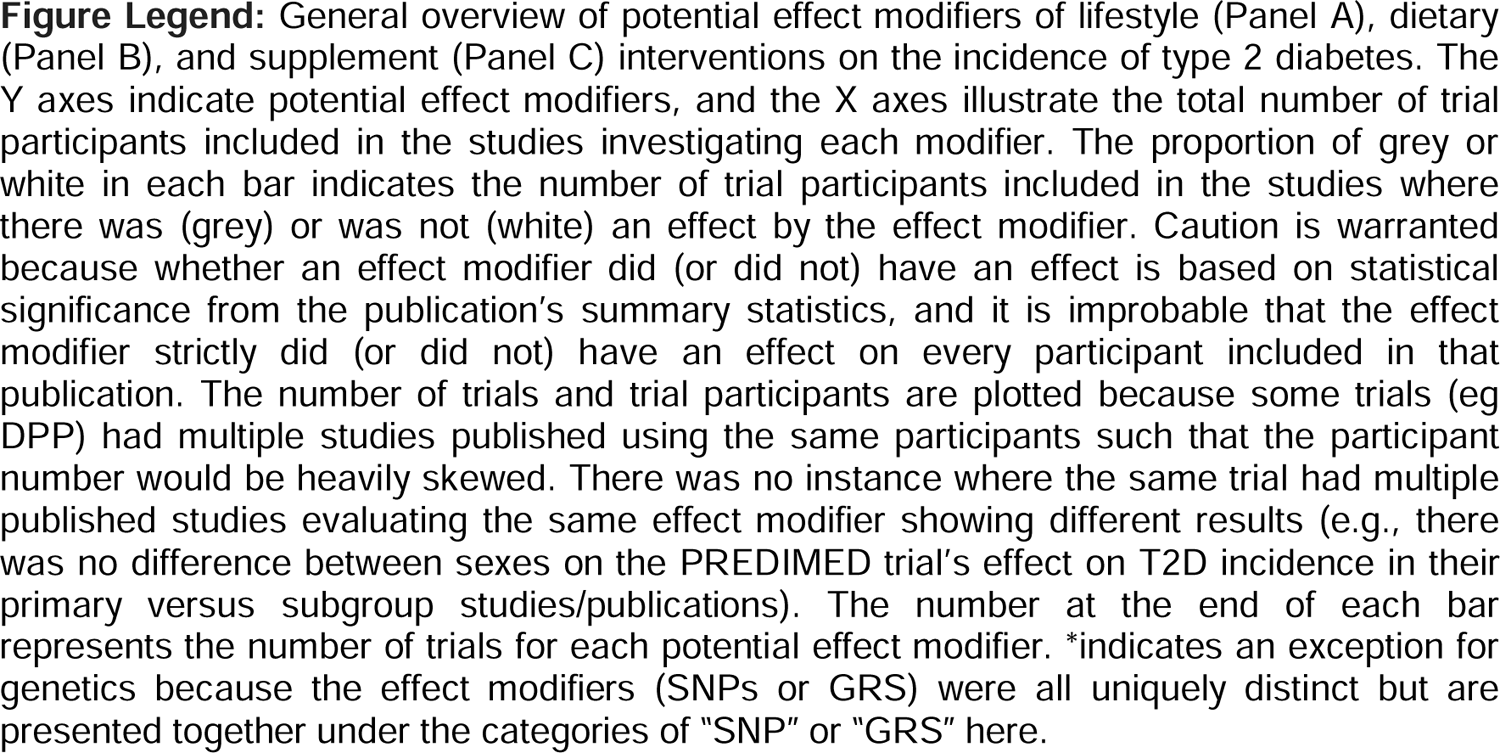
Potential effect modifiers of lifestyle, diet, and diet supplements intervention on the incidence of T2D

**Table 2:** Efficacy of T2D preventive interventions according to sociodemographic effect modifiers

|  | T2D preventive strategies |  |  |  |  |  |  |  |  |
| --- | --- | --- | --- | --- | --- | --- | --- | --- | --- |
|  | Intensive Lifestyle intervention |  |  | Dietary intervention |  |  | Dietary Supplements |  |  |
| Modifier | Number of Studies | Effect modification<sup>#</sup> | Certainty of evidence<sup>*</sup> | Number of Studies | Effect modification<sup>#</sup> | Certainty of evidence<sup>*</sup> | Number of Studies | Effect modification<sup>#</sup> | Certainty of evidence<sup>*</sup> |
| Age | 12 | Yes: 7 studies<br>No: 5 studies | Grade D | 3 | No: 3 studies | Grade D | 4 | Yes: 1 study<br>No: 3 studies | Grade D |
| Sex | 16 | Yes: 1 study<br>No: 15 studies | Grade D | 2 | No: 2 studies | Grade D | 1 | Yes: 1 study | Grade D |
| Race/ethnicity | 3 | No: 3 studies | Grade D | 1 | No: 1 study | Grade D | 1 | No: 1 study | Grade D |
| Socioeconomic status/<br>Education | 5 | Yes: 1 study<br>No: 4 studies | Grade D | - | - | - | - | - | - |
| Location | 2 | No: 2 studies | Grade D | - | - | - | 1 | No: 1 study | - |

**Table legend:** Overview of the included studies investigating whether sociodemographic factors modify the response to T2D preventive intervention strategies. <sup>#</sup> Yes/ No corresponds to significant/ nonsignificant effect modification, as reported in the study. <sup>\*</sup>Certainty of evidence denotes consistency, Grading based on Diabetes Canada scale A to D.

Fourteen studies investigated whether BMI modified the efficacy of multi-component lifestyle interventions. Nine of these studies showed that BMI is not associated with different responses to a lifestyle program, but five studies showed suggestive evidence that individuals with low BMI could benefit most from a lifestyle intervention. Among these five studies presenting evidence of the differential effect of a lifestyle intervention according to BMI, four were conducted in Asia (**Table 3**). No appreciable evidence for interactions with BMI was observed in studies that implemented a dietary or supplement intervention (**Table 3**). Eighteen studies tested the efficacy of an intensive lifestyle intervention for preventing T2D stratified based on baseline glucose levels, impaired glucose tolerance, or prediabetes status. Evidence presented in eight of these studies indicated statistically different effects based on baseline dysglycemia, but other studies did not find evidence of effect modifications. Three studies investigated family history of T2D as a potential lifestyle intervention effect modifier, and only one provided suggestive evidence of heterogenous treatment responses. Studies stratified by baseline cardiometabolic risk factors reported that individuals with poorer health status, particularly those with dyslipidemia and metabolic syndrome, tend to benefit more from dietary or supplement interventions than individuals who are healthier (**Table 3**).

**Table 3:** Efficacy of T2D preventive interventions according to clinical effect modifiers

|  | T2D preventive strategies |  |  |  |  |  |  |  |  |
| --- | --- | --- | --- | --- | --- | --- | --- | --- | --- |
|  | Intensive Lifestyle intervention |  |  | Dietary intervention |  |  | Dietary Supplements |  |  |
| Modifier | Number of Studies | Effect modification<sup>#</sup> | Certainty of evidence<sup>*</sup> | Number of Studies | Effect modification<sup>#</sup> | Certainty of evidence<sup>*</sup> | Number of Studies | Effect modification<sup>#</sup> | Certainty of evidence<sup>*</sup> |
| BMI | 14 | Yes: 5 studies<br>No: 9 studies | Grade D | 3 | No: 3 studies | Grade D | 4 | Yes: 1 study<br>No: 3 studies | Grade D |
| Prediabetes | 18 | Yes: 8 studies<br>No: 10 studies | Grade D | 1 | No: 1 study | Grade D | 1 | No: 1 study | Grade D |
| Family history | 3 | Yes: 1 study<br>No: 2 studies | Grade D | - | - |  | 3 | Yes: 2 studies<br>No: 1 study | Grade D |
| Dyslipidemia/<br>medications | 1 | No: 1 study | Grade D | 2 | Yes: 2 studies | Grade D | 2 | Yes: 1 study<br>No: 1 study | Grade D |
| Hypertension | - | - | - | 2 | Yes: 1 study<br>No: 1 study | Grade D | 2 | No: 2 studies | Grade D |
| Metabolic<br>syndrome | - | - | - | 1 | Yes: 1 study | Grade D | 1 | No: 1 study | Grade D |
| Menopausal<br>status, HRT<br>use | - | - | - | - | - | - | 3 | No: 3 studies | Grade D |

**Table legend:** Overview of the included studies investigating whether clinical factors modify the response to T2D preventive intervention strategies. <sup>#</sup> Yes/ No corresponds to significant/ nonsignificant effect modification, as reported in the study. <sup>\*</sup>Certainty of evidence denotes consistency, Grading based on Diabetes Canada scale A to D.

### Behavioral factors

Several secondary studies have assessed whether baseline lifestyle factors (i.e., overall dietary quality, alcohol intake, physical activity, and/or smoking) influence the efficacy of T2D prevention interventions. Evidence presented in studies investigating the effect of a lifestyle intervention according to baseline smoking status and physical activity indicates statistically different effects, suggesting that smokers and those with lower levels of physical activity benefited less from a lifestyle program (**Table 4**). Available studies reported no interactions of baseline smoking status and physical activity levels with dietary or supplement interventions on the risk of T2D. Among the four studies that focused on alcohol intake, only one found that the lifestyle intervention was more effective in individuals who drink alcohol frequently than in those who rarely drink. Six studies tested whether baseline diet modified the association between supplements and the risk of T2D and found no evidence of significant interactions (**Table 4**).

**Table 4:** Efficacy of T2D preventive interventions according to behavioral effect modifiers

|  | T2D preventive strategies |  |  |  |  |  |  |  |  |
| --- | --- | --- | --- | --- | --- | --- | --- | --- | --- |
|  | Intensive Lifestyle intervention |  |  | Dietary intervention |  |  | Dietary Supplements |  |  |
| Modifier | Number of Studies | Effect modification<sup>#</sup> | Certainty of evidence<sup>*</sup> | Number of Studies | Effect modification<sup>#</sup> | Certainty of evidence<sup>*</sup> | Number of Studies | Effect modification<sup>#</sup> | Certainty of evidence<sup>*</sup> |
| Smoking | 2 | Yes: 2 studies | Grade D | 1 | No: 1 study | Grade D | 3 | No: 3 studies | Grade D |
| Physical activity | 3 | Yes: 2 studies<br>No: 1 study | Grade D | 1 | No: 1 study | Grade D | 3 | No: 3 studies | Grade D |
| Alcohol intake | 1 | Yes: 1 study | Grade D | - | - | - | 3 | No: 3 studies | Grade D |
| Diet and supplements | - | - | - | 2 | No: 2 studies | Grade D | 6 | No: 6 studies | Grade D |

**Table legend:** Overview of the included studies investigating whether behavioral factors at baseline modify the response to T2D preventive intervention strategies. <sup>#</sup> Yes/ No corresponds to significant/ nonsignificant effect modification, as reported in the study. <sup>\*</sup>Certainty of evidence denotes consistency, Grading based on Diabetes Canada scale A to D.

### Molecular factors

The extent to which genetic predisposition modifies the efficacy of interventions to prevent T2D was reported in 21 publications, and most of them were based on data from the DPP and the DPS. Genetic predisposition was defined based on single genetic variants in 16 studies or genetic risk scores in five studies. While many of the T2D-associated loci identified in the earlier GWAS studies have been examined for their potential roles as effect modifiers, some reported evidence that individuals with specific genotypes could benefit the most from a lifestyle intervention, but these studies rarely corrected for the number of performed tests. Of the five studies that reported on the role of polygenic scores for T2D, only one study showed that lifestyle intervention was more effective among individuals with a high genetic risk.

Besides genetics, other molecular markers have also been studied for their potential effect modification of prevention interventions. A posthoc analysis in the PREDIMED trial showed that higher levels of plasma branched-chain amino acid levels at baseline attenuated the beneficial effect of a Mediterranean diet for the prevention of T2D, but these findings have been not replicated in independent studies. A separate study has found that individuals with low plasma levels of miR-29a, miR-28-3p, miR-126 and high plasma levels of miR-150 have a higher risk of developing T2D if they are allocated to a MedDiet, but not if they consume a low-fat and high-carbohydrate diet. (**Table 5**, **Figure 2**).

**Table 5:** Efficacy of T2D preventive interventions according to molecular effect modifiers

|  | T2D preventive strategies |  |  |  |  |  |
| --- | --- | --- | --- | --- | --- | --- |
|  | Intensive Lifestyle intervention |  |  | Dietary intervention |  |  |
| Modifier | Number of Studies | Effect modification<sup>#</sup> | Certainty of evidence<sup>*</sup> | Number of Studies | Effect modification<sup>#</sup> | Certainty of evidence<sup>*</sup> |
| T2D single SNPs | 16 | Yes: 10 studies<br>No: 6 studies | Grade D | 1 | Yes: 1 study | Grade D |
| Diabetes polygenic score | 5 | Yes: 1 study<br>No: 4 studies | Grade D | - | - | - |
| Metabolites/<br>miRNA | - | - | - | 3 | Yes: 3 studies | Grade D |

**Table legend:** Overview of the included studies investigating whether genetic and molecular factors at baseline modify the response to T2D preventive intervention strategies. <sup>#</sup> Yes/ No corresponds to significant/ nonsignificant effect modification, as reported in the study. <sup>\*</sup>Certainty of evidence denotes consistency, Grading based on Diabetes Canada scale A to D.

### Grading of evidence certainty

Although our systematic review included intervention studies, most of which were RCTs with low risk of bias, we evaluated certainty through our hypothesis of identifying valid effect modifiers to inform precision prevention. None of the studies included *a priori* consideration of intervention interactions with individual-level characteristics or risk factors in their study design, which were largely conducted as posthoc analyses. As a result, statistical power was often limited. Further, most did not adjust for individual-level risk factors, undermining validity of interpreting the role of effect modifiers independent of other traits. These considerations were factored into the major downgrading of the evidence (**Tables 2-5**)

## DISCUSSION

We performed a comprehensive systematic review to identify individual-level sociodemographic, clinical, behavioral, or molecular factors that could modify the efficacy of T2D prevention strategies. Overall, we find low to very low certainty of evidence that traits such as age, sex, BMI, race/ethnicity, socioeconomic status, baseline lifestyle factors, or genetics consistently and validly modify the effectiveness of lifestyle and behavioral interventions. Low-certainty evidence was observed for health status at baseline; individuals with poorer health, particularly those with prediabetes, were likely to benefit from prevention strategies compared to individuals in better health. However, whether the modest benefit reported in these studies was due to poor health status or other correlated risk factors cannot be ascertained based on the available evidence.

Large randomized clinical trials have consistently demonstrated that a healthy lifestyle or dietary interventions can prevent or delay T2D.^3, 4, 6, 15^ However, there is large inter-individual variability in response to these preventive interventions, in which some people seem to greatly benefit from T2D preventive interventions. Precision prevention aims to identify participant characteristics that determine this variability in response to ultimately tailor preventive strategies to subgroups of individuals that are likely to benefit the most. So far, no studies exist that were prospectively designed to determine interactions by a baseline trait or factor with an intervention in prevention of T2D. We evaluated the evidence base and identified several stratified posthoc analyses of existing prevention intervention trials. In posthoc analyses, the participant population is stratified by a potential effect modifier, and the efficacy of the intervention is tested within each stratum and compared across the strata, which reduces statistical power and increases type 2 error. Furthermore, precision prevention strategies may be optimized by incorporating several individual-level factors into decision making, whereas the current literature is predominantly evaluating one stratified trait as a time. For example, correlated behaviors such as physical activity, diet, and smoking, might provide more information when considered collectively than on their own. Clinical trials specifically designed to investigate the influence of sociodemographic, clinical, behavioral, or molecular factors on the response to T2D preventive strategies are needed to generate valid and robust evidence before the implementation of T2D precision prevention strategies.

One area of promise warranting further research is the presence of prediabetes at baseline and whether this may be targeted in future precision prevention research. Low certainty evidence suggests that individuals at risk of T2D or with prediabetes at baseline benefit slightly more of prevention interventions than individuals who are not at risk of T2D.^3–6^ However, the evidence is inconsistent, even though the studies report that a lifestyle intervention, compared to standard care, results in higher T2D reduction rates among studies conducted in Asia.^15–18^ Beyond the methodological limitations of the available evidence, an additional reason for inconsistent evidence supporting the greater effectiveness of lifestyle interventions for the prevention of T2D among individuals with prediabetes is due to the heterogeneity that characterizes this condition. Prediabetes refers to a pathophysiological state of early alterations in glucose metabolism that precedes the development of diabetes, but the mechanism by which glucose is elevated are very different and could range from those with primary alterations in insulin secretion pathways to those with primary insulin resistance.^19^ Clinical trials specifically designed to capture the nuances and complexity of early glycemic alterations and whether individuals with distinct pathophysiological features benefit from more targeted preventive interventions are needed to fill the gap in current T2D precision prevention evidence.

Even though there are far more lifestyle intervention trials for the prevention of T2D than diet alone and diet supplementation trials, collectively, however, results for effect modification by any one factor are sparsely reported or arising from an evidence base of very different trials and patient populations. Further, many of the secondary analyses included in this systematic review are derived from two single clinical interventions. Findings from available evidence contrast with recent clinical studies documenting variable responses to identical foods, diets, or lifestyle interventions based on inter-individual differences in demographic, clinical, genetic, gut microbiota, and lifestyle characteristics.^20–22^ While these studies offer insights into variable postprandial metabolic response, their short follow-up periods, the lack of time-series data and changes in parameters that could influence response to interventions, and the inclusion of relatively young and healthy individuals preclude the generalizability to T2D prevention efforts. Whether the promise of T2D precision prevention is matched by evidence of the long-term beneficial impact remains uncertain. Still, interest and activity in this field are proliferating to identify factors underlying variable nutritional responses and develop algorithms to predict individual responses to nutrients, foods, and dietary patterns.

Our systematic review had some limitations. The scope of our literature review as part of the PDMI was broad and inclusive of diverse study designs, T2D prevention strategies, study populations, and effect modification analyses. Although this resulted in a heterogeneous evidence base and did not provide an opportunity for meta-analysis, we qualitatively synthesized the evidence for precision prevention generally. Our hypothesis originally spanned to include observational studies, which were ultimately excluded due to the uncertainty of their being readily related to clinical interventions. Protocol amendments were registered to reflect these decisions prior to study screening and extraction.

In conclusion, our systematic review and synthesis of the T2D prevention literature provide low to very low certainty evidence that sociodemographic, clinical, lifestyle, or molecular factors are more useful, valid, and consistent in informing T2D precision prevention strategies than current interventions. We also uncover several areas of potential for growth in the precision medicine field, including prospectively designed interventions and clinical trials incorporating the investigation of treatment response heterogeneity.

## Data Availability

All data produced in the present work are contained in the manuscript

## AUTHOR CONTRIBUTIONS

DB, RM, SLF, JSP, PWF, DT, JM, VM, and RJFL contributed to the conception and design of the research questions. All authors contributed to the study screening and data extraction. DT and JM did the quality assessment; DB, RM, VS, MN, SLF, MGF, JSP, MRL, DT, JM, VM, and RJFL summarized and interpreted the data. DB, JM, and RJFL drafted the paper; DT and VM revised it substantively. All authors edited the manuscript and approved the final version.

## Acknowledgment

We thank Hugo Fitipaldi, Esther González-Padilla, Alisha Sha, and Jiaxi Yang for attending some of the working group meetings and/or for reviewing some of the abstracts.

The *Precision Medicine in Diabetes Initiative* (PMDI) was established in 2018 by the American Diabetes Association (ADA) in partnership with the European Association for the Study of Diabetes (EASD). The ADA/EASD PMDI includes global thought leaders in precision diabetes medicine who are working to address the burgeoning need for better diabetes prevention and care through precision medicine [Nolan et al, Diabetes Care, 2022]. This Systematic Review is written on behalf of the ADA/EASD PMDI as part of a comprehensive evidence evaluation in support of the 2^nd^ International Consensus Report on Precision Diabetes Medicine [Tobias et al, Nat Med year?].

