## Supplement for "Role of sociodemographic, clinical, behavioral, and molecular factors in precision prevention of type 2 diabetes: a systematic review"

**Table S1:** Search terms used for retrieving the studies of interest

(2000/01/01:2021/07/15[Date - Publication] AND ("diabetes mellitus, type 2"[MeSH Terms] OR "diabetes type-2"[Title/Abstract] OR "diabetes type-2"[Title/Abstract] OR "T2D"[Title/Abstract] OR "type 2-diabetes"[Title/Abstract] OR "type 2-diabetes"[Title/Abstract] OR "diabetes type-ii"[Title/Abstract] OR "diabetes type-ii"[Title/Abstract] OR ("diabetes mellitus, type 2"[MeSH Terms] OR "type 2 diabetes mellitus"[All Fields] OR "type 2-diabetes"[All Fields])) AND ("weight loss"[MeSH Terms] OR "weight reduction programs"[MeSH Terms] OR "diet, reducing"[MeSH Terms] OR "body weight*"[Title/Abstract] OR "bodyweight*"[Title/Abstract] OR "body composition*"[Title/Abstract] OR "waist circumferen*"[Title/Abstract] OR "body fat"[Title/Abstract] OR "weight loss"[All Fields] OR "weight change"[All Fields] OR "bmi"[All Fields] OR "body mass"[All Fields] OR "diet"[MeSH Terms] OR "dietary supplements"[MeSH Terms] OR "diet, carbohydrate restricted"[MeSH Terms] OR "diet, fat restricted"[MeSH Terms] OR "caloric restriction"[MeSH Terms] OR "diet"[Title/Abstract] OR "food"[Title/Abstract] OR "diet therapy"[Title/Abstract] OR "diet intervention*"[Title/Abstract] OR "macronutrient"[Title/Abstract] OR "nutrition"[Title/Abstract] OR "diet*"[Title/Abstract] OR "supplement*"[Title/Abstract] OR "protein*"[All Fields] OR "meat"[All Fields] OR "food*"[All Fields] OR "beverage*"[All Fields] OR "meal*"[All Fields] OR "exercise"[MeSH Terms] OR "physical fitness"[MeSH Terms] OR "life style"[MeSH Terms] OR "healthy lifestyle"[MeSH Terms] OR "sedentary behavior"[MeSH Terms] OR "exercise"[Title/Abstract] OR "physical activity"[Title/Abstract] OR "fitness"[Title/Abstract] OR "sedentary"[Title/Abstract] OR "walk*"[Title/Abstract] OR "stretch*"[Title/Abstract] OR "lifestyle"[Title/Abstract] OR "strength train*"[Title/Abstract] OR "wellness"[Title/Abstract]) AND (("primary prevention"[MeSH Terms] OR "prevent*"[Title/Abstract] OR "inciden*"[All Fields] OR ("prevent"[All Fields] OR "preventability"[All Fields] OR "preventable"[All Fields] OR "preventative"[All Fields] OR "preventatively"[All Fields] OR "preventatives"[All Fields] OR "prevented"[All Fields] OR "preventing"[All Fields] OR "prevention and control"[MeSH Subheading] OR ("prevention"[All Fields] AND "control"[All Fields]) OR "prevention and control"[All Fields] OR "prevention"[All Fields] OR "prevention s"[All Fields] OR "preventions"[All Fields] OR "preventive"[All Fields] OR "preventively"[All Fields] OR "preventives"[All Fields] OR "prevents"[All Fields])) AND ("cohort studies"[MeSH Terms] OR "clinical study"[Publication Type] OR "cohort"[Title/Abstract] OR "longitudinal"[Title/Abstract] OR "prospective"[Title/Abstract] OR "intervention"[Title/Abstract]))) NOT ("letter"[Publication Type] OR "review"[Publication Type] OR "editorial"[Publication Type] OR "comment"[Publication Type] OR "models, animal"[MeSH Terms] OR "in vitro techniques"[MeSH Terms])

**Table S2:** Template used for extraction of data

| **Study Level Information** | PMID |
| --- | --- |
|  | Trial/Study Name |
|  | Clinical Trial Registry # |
|  | Continent |
|  | Country |
| **Study Design** | Treatment groups (Parallel/ cross-over/ single arm, Not reported) |
|  | Assignment level (Individual, cluster, Not reported) |
|  | Randomization (Yes/ No/ Not Reported) |
|  | Enrollment year(s) |
|  | Enrollment N |
| **Study Population** | Population (General population/ prediabetes/ overweight/ obese/ Others) |
|  | Age range |
|  | BMI range |
| **Intervention Information** | Total treatment groups |
|  | Interventions (Placebo/ Standard Care/ Multi component Lifestyle/ Diet/ Diet Supplements/ Mental health/ Physical activity/ weight loss/smoking cessation/ sleep/ Others) |
| **Treatment groups description** | Treatment name |
|  | Brief description |
|  | Baseline N |
| **Outcome Information** | T2D ascertainment (Self-reported/ In person OGTT/ healthcare Registry) |
|  | Diagnostic criteria |
| **Results** | Study end date |
|  | Follow-up: (mean/ median/ range/SD) |
|  | Follow-up: unit |
|  | T2D cases (n) |
| **Results by sub-groups/strata** | Subgroup categories: |
|  | 1. Social/environment strata: (Race/ethnicity, Income, Education, Built environment, Insurance status, SES score) |
|  | 2. Demographics strata: (Age, Sex, family history of diabetes) |
|  | 3. Clinical trait/health status strata: (BMI, Hip, waist, other anthropometric, Prediabetes, Glycemic trait/severity, Dyslipidemia, Hypertension) |
|  | 4. Genetics and omics: (single SNPs, Genetic Risk Score, Metabolomics, Proteomics, Microbiome) |
| **Strata-specific Results** | Stratified variable |
|  | Number of strata |
|  | List strata |
|  | Absolute treatment effect (T2D cases, IR, cumulative incidence, NNT, etc.) |
|  | Relative treatment effect (RR, OR, RRR, IRR, etc.) |
|  | P-value effect modification |
|  | Data not shown: "significant effect modification" |
|  | Data not shown: "no effect modification" |
|  | Location of results |

**Table S3.** Additional information on included studies

| **Trial/Study Name** | **Primary Outcome(s) of Original Intervention** | **T2D Follow-up Duration - Years** | **Main Effect on T2D Risk** |
| --- | --- | --- | --- |
| CORDIOPREV | CVD | NR | NR |
| PREDIMED | CVD (MI, stroke, CVD death) | Median 4.1 years | Mediterranean + EVOO HR=0.64 (95% CI=0.44, 0.94) vs. Low-Fat; Mediterranean + Nuts HR=0.87 (0.63, 1.20) vs. Low-Fat |
| Shahbazi 2018 | T2D | 2 years | High fat diet vs. Standard diet RR=0.43 (95% CI=0.1, 0.9), p=0.03; Normal fat diet vs. Standard diet RR=0.60 (0.2, 1.2), p=0.1 |
| WHI-DM | Breast and colorectal cancers | Median 17.3 years | Intervention period: HR=0.96 (95% CI=0.90, 1.03); Intervention + Observation periods: HR=0.96 (0.91, 1.00) |
| ATBC | Lung cancer | Mean 12.5 years | Alpha-tocopherol vs. Placebo RR=0.91 (95% CI=0.73, 1.12); Beta-carotene vs. Placebo RR=0.97 (0.79, 1.20); Alpha-tocopherol + beta-carotene vs. Placebo RR=0.91 (0.73, 1.12) |
| D2d | T2D | Median 2.5 years | HR=0.88 (95% CI=0.75, 1.04), p=0.12 |
| WAFACS | Secondary CVD | Median 7.3 years | HR=0.91 (95% CI=0.76, 1.09) |
| WACS | Secondary CVD | Median 9.2 years | Vitamin C vs. Placebo RR=0.89 (95% CI=0.78, 1.02), p=0.09; Vitamin E vs. Placebo RR=1.13 (0.99, 1.29), p=0.07; Beta-carotene vs. Placebo RR=0.97 (0.85, 1.11), p=0.68 |
| WHS | CVD and cancer | Mean 10.0 years | RR=0.95 (0.87, 1.05), p=0.31 |
| Chae 2012 | Not stated | 2 years | Adjusted OR=0.77 (95% CI=0.60, 1.00), p=0.038 |
| Da Qing IGT and Diabetes Study | T2D | 6 years | IR (cases/100 PY): Diet=10.0 (95% CI=7.5,12.5) vs. Exercise=8.3 (6.4, 10.3) vs. Diet + Exercise=9.6 (7.2, 12.0) vs. Control=15.7 (12.7, 18.7); p<0.05 for all interventions vs. Control |
| D-CLIP | T2D | 3 years | HR=0.68 (95% CI=0.50, 0.93) |
| DE-PLAN-CAT | T2D | Median 4.2 years | HR=0.64 (95% CI=0.47, 0.87), p=0.004 |
| DPP | T2D | Mean 2.8 years | Lifestyle vs. Placebo HR=0.42 (95% CI=0.34, 0.52); Metformin vs. Placebo HR=0.69 (0.57, 0.83); Lifestyle vs. Metformin HR=0.61 (0.49, 0.76) |
| EDIPS-Newcastle | T2D | Mean 3.11 years | RR=0.45 (95% CI=0.2, 1.2) |
| Finnish DPS | T2D | Mean 3.2 years | HR=0.4 (95% CI=0.3 to 0.7), p<0.001 |
| IDPP-2013 | T2D | Mean 20.2 months | HR=0.640 (95% CI=0.446, 0.917), p=0.015 |
| IDPP-1 | T2D | Median 30 months | Lifestyle vs. Control HR=0.715 (95% CI=0.627, 0.795); Metformin vs. Control HR=0.736 (0.649, 0.809); Lifestyle + Metformin HR=0.718 (0.630, 0.797) |
| IDPP-2 | T2D | 36 months | HR=0.984 (95% CI=0.672, 1.443), p=0.936 |
| Japan DPP | T2D | Mean 2.3 years | IR (cases/100 PY) Lifestyle=2.7 vs. Control=5.1 |
| K-DPP | T2D | 24 months | RR=0.88 (95% CI 0.66–1.16); p = 0.36 |
| Kosaka 2004 | T2D | 4 years | Cumulative incidence: Lifestyle=3.0% vs. Control=9.3%; p<0.001 |
| Let's Prevent Diabetes | T2D | 36 months | HR=0.74 (95% CI=0.48, 1.14); p=0.18 |
| MRFIT | CHD death | 6 years | HR=1.08 (95% CI=0.96, 1.20) |
| Nanditha 2020 | T2D | 24 months | HR=0.89 (95% CI=0.74, 1.07), p=0.22 |
| FIN-D2D | T2D | -- | -- |
| NMB-Trial | T2D | 3 months | RR=0.362 (0.302, 0.435) |
| NDPS | T2D | 24.6 months | Lifestyle vs. Control HR=0.53 (96% CI=0.35, 0.81), p=0.003; Lifestyle + Peers vs. Control HR=0.64 (0.43, 0.97), p=0.03 |
| PreDE | T2D | 24 months | RR=0.68 (0.47, 0.99) |
| TLGS | Non-communicable diseases | 3.6 years (SD=1.0) | HR=0.35 (95% CI=0.17, 0.70); p=0.003 |
| Thai DPP | T2D | 24 months | IR (cases/1,000 PY): Lifestyle=12.1 (95% CI=10.7, 13.8) vs. Control=16.6 (95% CI=14.6, 18.8); p<0.001); HR=0.71 (95% CI=0.59, 0.85) |
| VIP | CVD, T2D | -- | -- |
| Zensharen Study | T2D | Mean 32.1 months | HR=0.56 (95% CI=0.36-0.87) |

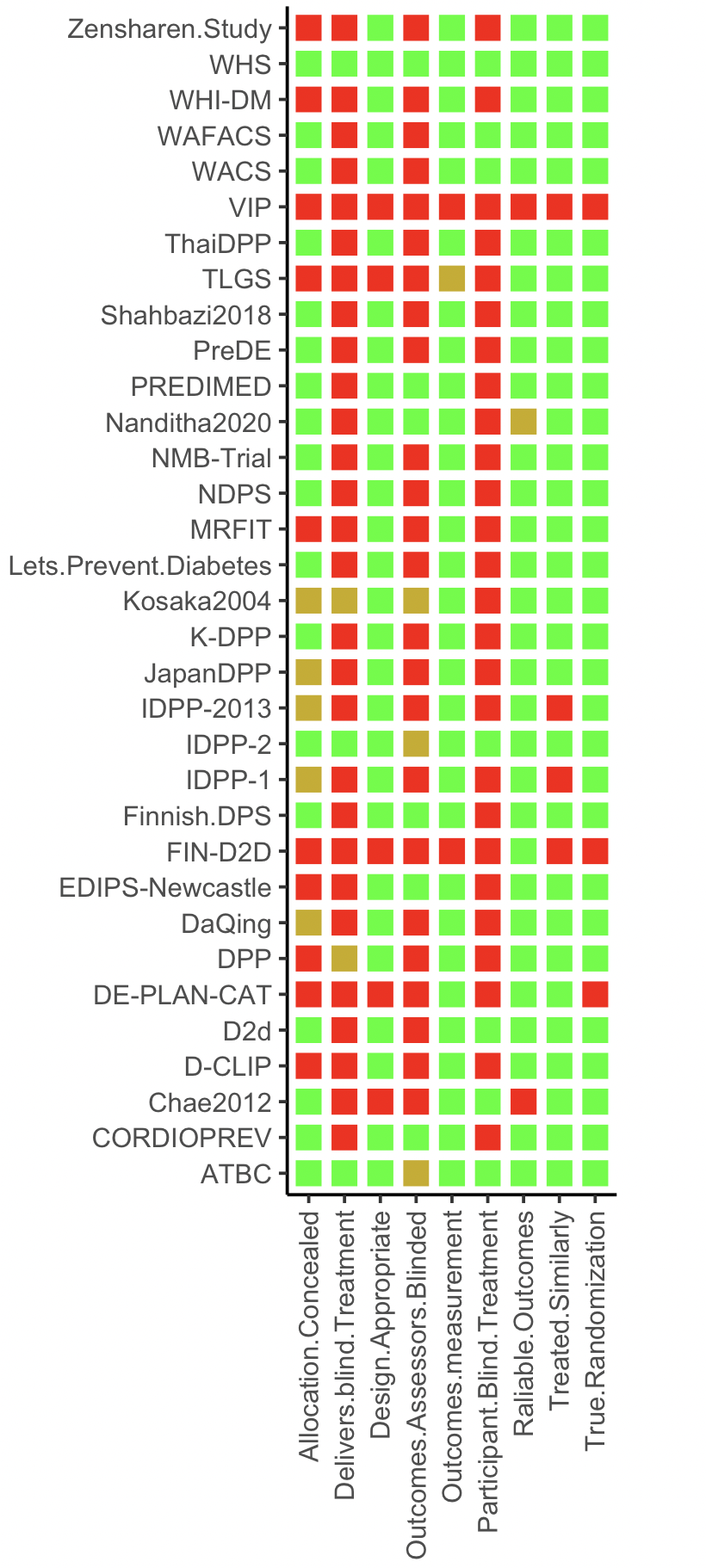
**Figure S1:** Trial-level quality assessment

**Figure Legend:** We used 9 questions from the JBI Critical Appraisal Checklist tailored to evaluating the quality of the study design with consideration for our primary interest in stratified results rather than the total intervention effect for T2D risk. “Yes” is represented in green color, “No” in red color, and “Not available/Unknow” in yellow color.

**Figure S2:** Sample size and the duration of intervention across the trials

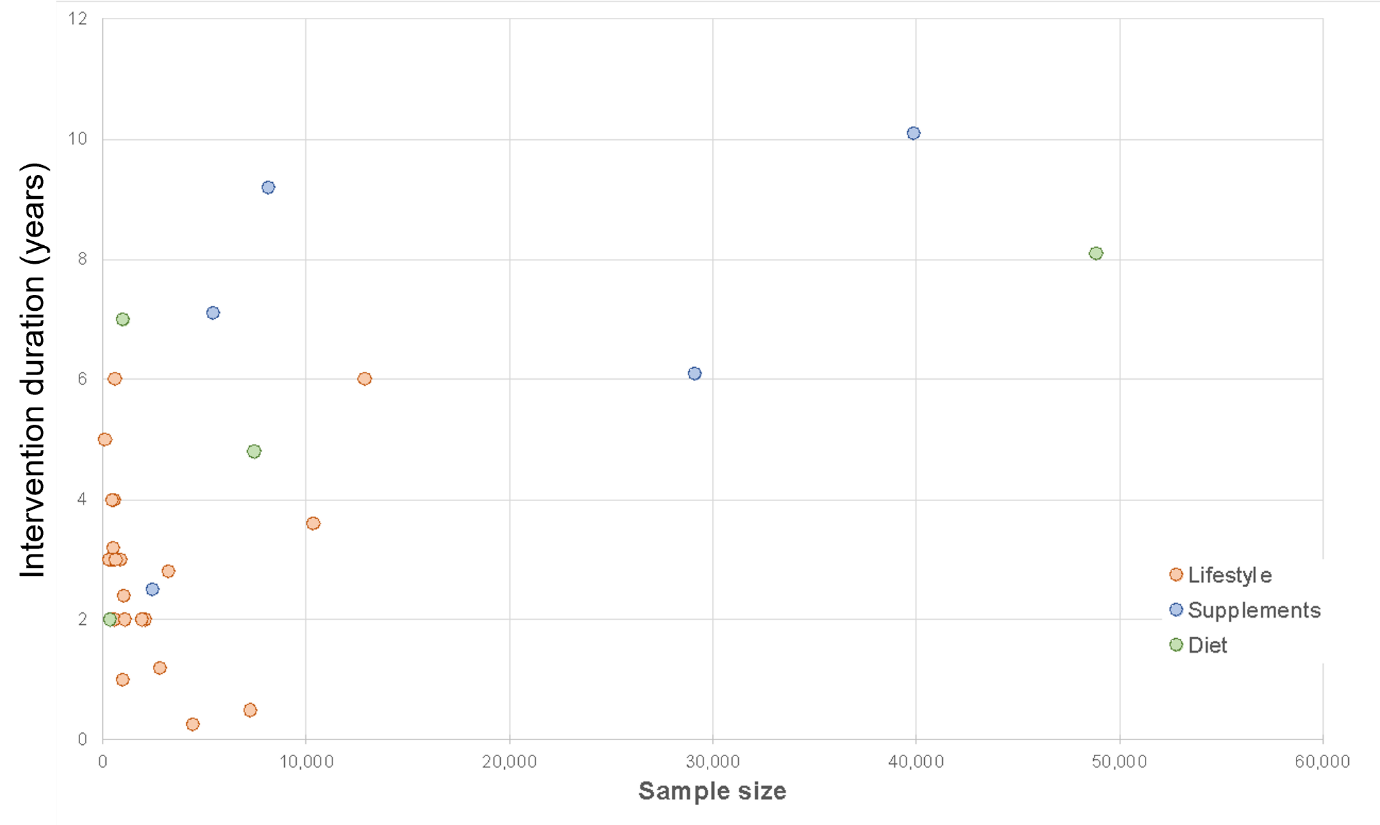

**Figure Legend:** Scatterplot to show the distribution of sample size and the intervention years of the completed trials. The Västerbotten Intervention Programme (VIP) with more than 113,000 individuals and ongoing since 1985 has not been included in the above plot.
